# Statistical segmentation and correlation analysis of the EEG between the different phases of general anesthesia

**DOI:** 10.1101/2022.05.27.22275606

**Authors:** C. Sun, D. Longrois, D. Holcman

## Abstract

Electroencephalography (EEG) is routinely used to monitor general anesthesia (GA). Unanswered questions concern the possibility of using the EEG signal to classify patients as more or less sensitive to anesthetic drugs from the onset of anesthesia. We investigate here possible statistical correlation between different phases of general anesthesia. We test whether it could be possible to predict the speed of return to consciousness from the induction or the maintenance phases. For that goal, we tracked the maximum power of the *α*−band and follow its time course. Using an optimization procedure, we quantify the characteristic shift of the *α*−band during recovery and the associated duration. Interestingly, we found no correlation (Pearson coefficient) between these shifts and the amount of *α*−band or iso-electric suppressions (flat EEG epochs) present in the maintenance phase or induced by a propofol injection during induction. To quantify the instability of the *α*−band, we introduce the total variation the *α*−band that accounts for all possible deviation from a flat line. To conclude, the present analysis shows that it would not be possible to anticipate the duration of recovery of consciousness from previous phases of general anesthesia in children and adults. Possibly the involved neuronal mechanisms during the different phases are not comparable.

## 1 Introduction

General anesthesia (GA) can be decomposed into three phases: induction, maintenance and recovery. Monitoring GA is at present based on frontal cortical electroencephalogram (EEG) features extraction in real time. The main interest of EEG-based GA is to avoid deep sedation or cortical awareness. Such monitoring allows the anesthesiologist to define the adequate hypnotic dose to inject. Yet, optimizing the brain monitoring remains a challenge in the absence of any optimization principle [1, 2, 3, 4, 5]. Prevention of anesthetic drugs overdose as opposed to correction after diagnosis would probably reduces postoperative complications such as delirium, cognitive dysfunction that were shown to be associated with the presence of iso-electric suppressions (IES) [6, 7].

Injection of an anesthetic agents such as propofol or inhalation of sevoflurane induces a change in the brain activity leading to the emergence of dominant α−oscillations in the (8-12) Hz frequency band. At the end of anesthesia, the hypnotic is turned off and a spectral analysis reveals a stereotypical zip-shape signature where the α−band increases during recovery of consciousness (ROC) while the *δ*− band decreases before disappearing [8, 9]. The EEG patterns during ROC have received much less attention than the maintenance phase. In both children and adults patients, it is necessary to define the adequate moment for extubation. Too early or late extubation would increase the risk of respiratory complications [10]. Furthermore, the speed of ROC would also change the way emergency from GA is managed. Therefore, the question arises whether information derived from the EEG signal during induction and maintenance phases could predict the patterns of recovery from GA. Specifically, it remains unclear whether there are correlation indices between the duration and the frequency shift of the α−band during the transient increase associated to ROC. These correlations could be relevant to anticipate the recovery time scale from the previous stages of anesthesia.

To study correlations between the EEG patterns associated with the different phases of GA, various mathematical parameters can be extracted from the filtered EEG. These parameters are for example the fraction of IES or α−band suppressions, computed by filtering the EEG signal with wavelet transform [11, 12, 13, 14, 15]. Several adaptative methods are now used to analyse, to correct [16, 17, 18, 19, 20] and to extract fundamental features from the EEG signal in real-time such as wavelet transform, independent and principal component analysis or diffusion-based classification [21, 22]. We shall use here the total variation *V*_*α*_ of the maximum *α*−frequencies located in the EEG signal [23], which quantifies the deviation of *α*−oscillations maximum amplitude from a straight line. We also estimate from a fitting procedure, the duration of the recovery, during which the frequency of the *α*−oscillations shifts significantly after the hypnotic injection or inhalation is stopped. We use these parameters to study the possible correlations between the ROC and the previous phases of anesthesia. To define the duration and the frequency shifts of the recovery phase, we fit the power of the *α*−band with model functions.

To detect correlations between the α−band dynamics the power, as well as the amount of iso-electric suppressions, we develop various segmentation of the EEG signal and perform a statistical analysis in cohorts of adults and children. Our hypothesis is that EEG features consistent with overdosing of anesthetic drugs could lead to an increase in the time of ROC. As we shall see below, we report in fact no correlation between the main statistical parameters extracted from the maintenance or the induction phases of GA and the recovery phase, confirming some of the conclusion of a previous study based on different markers [24]. The present analysis suggests that different brain mechanisms could be involved during ROC and that it is not possible from the EEG signal alone to anticipate the properties of the ROC.

## 2 Materials and Methods

### 2.1 Statistical analysis

We used MATLAB R2021a software to perform the statistical analysis. The significance level used in this study was *α*=0.05. The values were expressed in percentage for qualitative variables and median (Inter-Quartile Range: [IQR]) for quantitative variables. Variables were correlated using Pearson correlation test.

### 2.2 EEG recordings and preprocessing

The EEG monitoring was done by the Masimo Sedline^®^ monitor with four frontal electrodes F7, Fp1, Fp2, F8 and a sampling frequency of 173 Hz.

### 2.3 Detection of two reference points defining the recovery phase

To detect the recovery phase of the EEG, we introduced two reference time points *t*_*out*_ and *t*_*in*_ to characterize how the EEG evolves during the recovery phase. The method is based on following the decrease of the *α*−band power *P*_*α*_ as well as the increase of the maximum power frequency *f*_*max*_ towards *β* rhythms.

We first fit the *α*−band power *P*_*α*_ by a sigmoid function 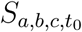 defined by

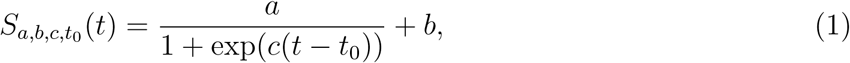

where the parameters *a, b, c* and *t*_0_ are estimated by a minimization procedure. We define the minimization routine to estimate the parameters:

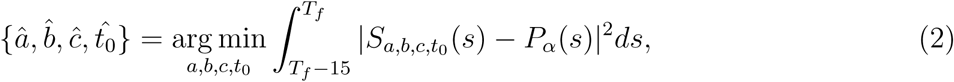

where *T*_*f*_ is the time where the EEG recordings terminates. We define *t*_*out*_ time point as the first time the sigmoid curve of 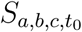 crosses the constant curve *y* = *aT* + *b* where the threshold *T* = 0.1. In practice, we have to solve 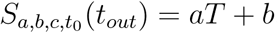, thus

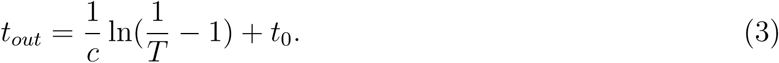

We thus fit *f*_*max*_ with an exponential curve 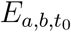 defined by

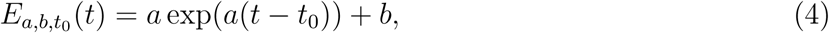

where *a, b* and *t*_0_ are estimated by

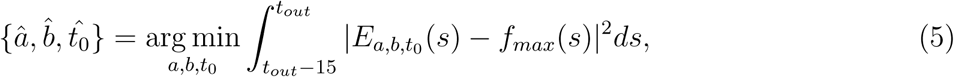

We define time point *t*_*in*_ as the first time where the exponential curve 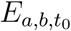 crosses the line *y* = *aT* + *b*, where the threshold 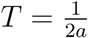. In practice, we search for the time point where 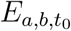 deviates by 0.5 Hz from the asymptotic line *y* = *b*. Finally, we chose

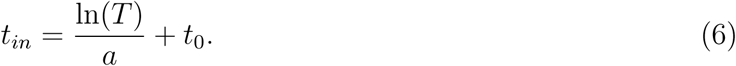

### 2.4 Segmenting the iso-electric suppressions

We now describe the segmentation procedure of the IES: we first apply a bandpass filter to the EEG signal *S*(*t*) in the range [8,16] Hz leading to the output filtered signal *S*_*α*_(*t*). We then normalize *S*_*α*_(*t*) by its Root Mean Square (RMS) to get

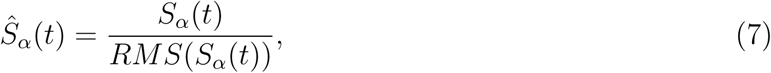

where

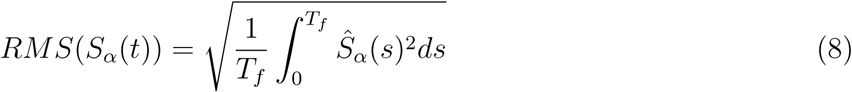

We then estimate the upper *S*_*up*_(*t*) (resp. *Ŝ*_*α,up*_(*t*)) and lower *S*_*low*_(*t*) (resp. *Ŝ*_*α,low*_ (*t*)) envelops of *S*(*t*) (resp. *Ŝ*_*α*_(*t*)) by interpolating the local maxima and minima. We compute their differences:

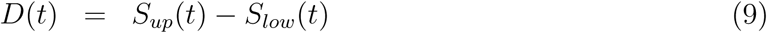

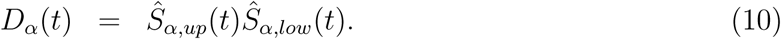

We define two IES threshold values

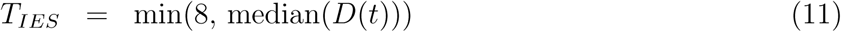

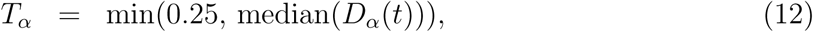

and we look for the set of time points satisfying the conditions:

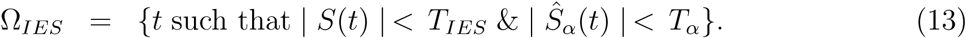

We then aggregate the time points of Ω_*IES*_ into time segments such that each pair of consecutive points of the segment matches with the sampling frequency which is known. To smooth the effect of the hard thresholding in Ω_*IES*_, we use mathematical morphological erosion and dilation [26] to gather the segments together with the conditions that the maximum separation length do not exceed 0.9 second while the minimum length of each segment is at least 1.1 second. After this morphological procedure, we obtain an ensemble of IES times where the EEG signal is of amplitude less than *T*_*IES*_ and its normalized signal in the high frequency domain is of amplitude less than *T*_*α*_.

### 2.5 Mathematical indicators associated to the iso-electric suppression and the *α*−band

- The total time spent in IES is computed by taking the sum of all time segments in Ω_*IES*_. For each i-th segment which starts (resp. ends) at time *T*_*i,start*_ (resp. *T*_*i,end*_), we define

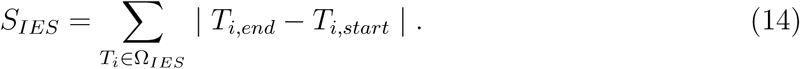
- The longest IES event is computed as the maximum of the duration present in Ω_*IES*_.

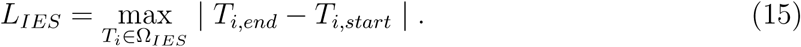
- The *α*−power 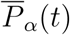 at time t describes the energy of the signal in the range [8, 12] Hz.

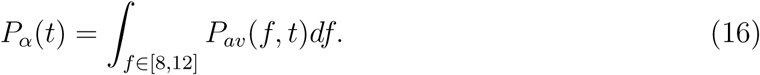 We define the mean value of the α−band power during the maintenance phase as the approximation of the discretized sum

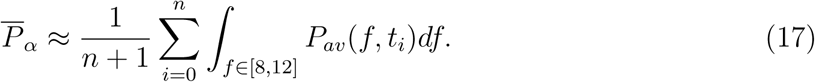
- The maximum power frequency *α*_*max*_ within the *α*−band is the frequency for which the power is maximal. It is computed as the frequency where the average power *P*_*av*_(*f, t*) reaches its maximum for time *t* for any frequency in the *α*−band range.

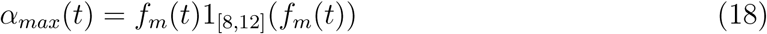

where

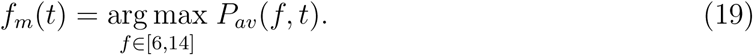 Due to the construction of the function *α*_*max*_(*t*), it contains oscillations at multiscale, that do not necessarily reflect physiological changes. We thus decided to smooth *α*_*max*_(*t*) using the Savitzky-Golay smoothing filter [27] to keep lower frequencies (Fig.1). We use a sliding window of 2 minutes size and 5 seconds step size with order 1 polynomial regression. The resulting filtered signal is 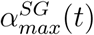.
- The total variation *V*_*α*_ of the function 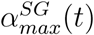 counts the cumulated amplitude of all local oscillations and thus measures the deviation of the maximum α−band frequency dynamics from a flat line. It is well known measure in the theory of function [28, 23]. Thus a large *V*_*α*_ characterized a global instability during anesthesia. In practice, when a time discretization (*t*_*i*_, ..*t*_*n*_) is chosen, it is computed as the sum of the absolute difference between two consecutive frequency time points, as follows:

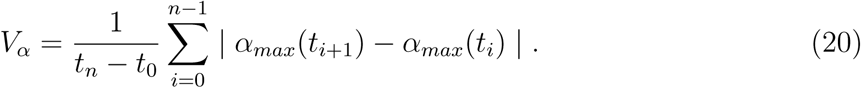 This computation is equivalent to estimating the differences at local maxima *a*_*i*_ and minima *b*_*i*_ time points over the duration *T*:

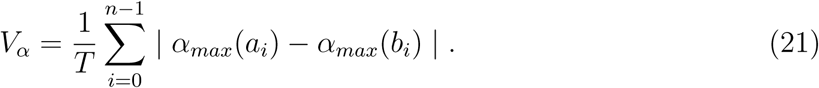

**Figure 1:**
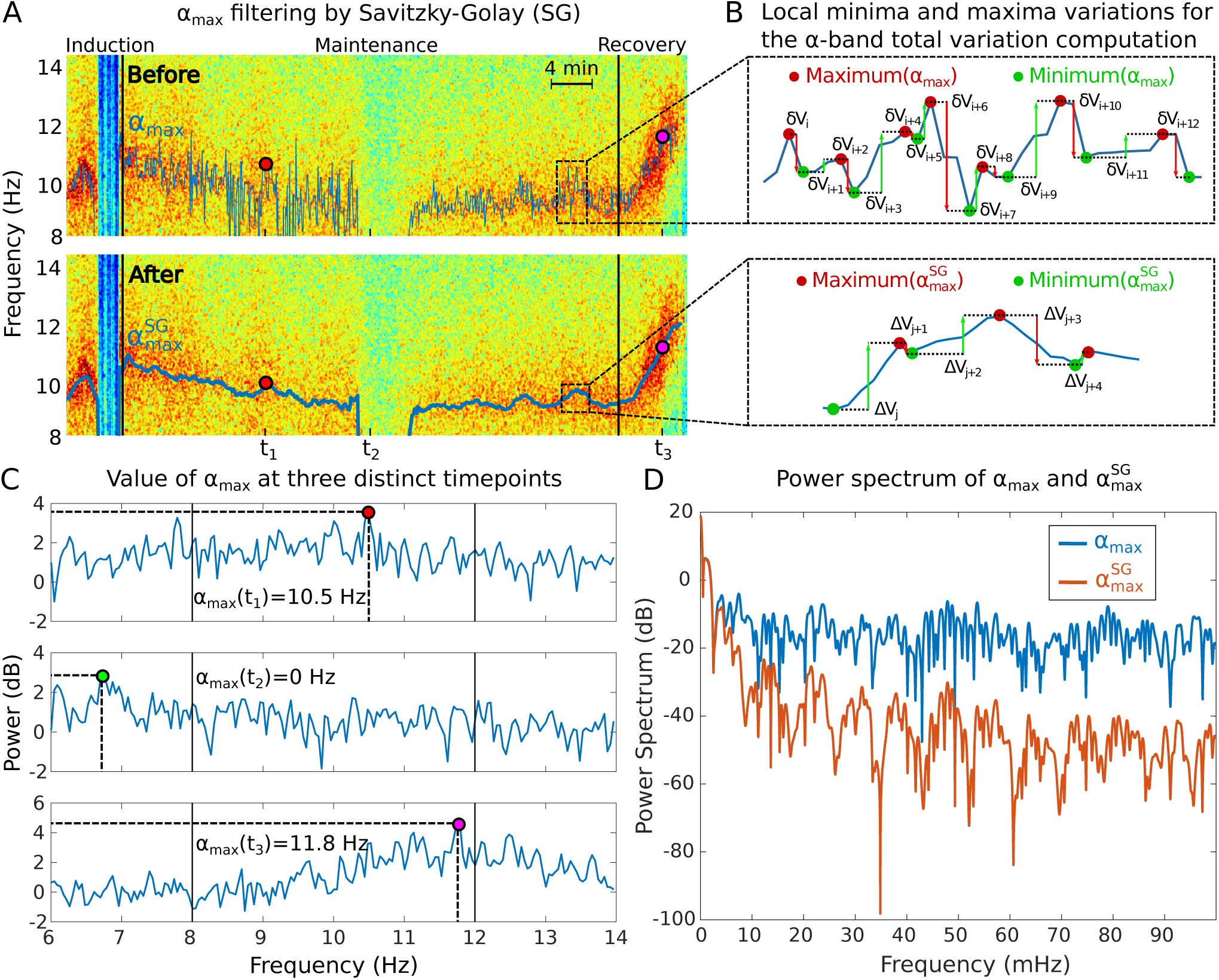
Total variation *V*_*α*_ analysis associated to the *α*−band. **(A)** Maximum power frequency *α*_*max*_ of the *α*−oscillations before (top) and after (bottom) applying the Savitzky-Golay (SG) filter to remove the high frequencies. **(B)** Reduction of the local minima and maxima variations of the *α*_*max*_ from the initial signal(top) to the filtered one (bottom). **(C)** Illustration of *α*_*max*_ computation for three time points (*t*_1_, *t*_2_ and *t*_3_): when the maximum power frequency is outside the *α* range, the value of *α*_*max*_ is set to zero, otherwise it is equal to the maximum power frequency value. **(D)** Power spectra of *α*_*max*_ before (blue) and after (red) SG filtering showing that only the slowest oscillations remain in the filtered signal.

## 3 Results

During the maintenance phase of GA, the frequency spectrum of the EEG signal tends to be dominated by the presence of two main bands: the *δ* band (0.1-4) Hz and α−band (8-12) Hz, as shown in Fig.2A-B-C. We first describe the segmentation of the recovery phase, where we estimate the shifts in time and frequency associated with this recovery upon an optimization procedure. We then use these parameters to find possible correlations between them and the total time spent in suppression (iso-electric EEG), a marker of brain sensitivity [14]. We further introduce the longest suppression induced by a propofol bolusupon induction of GA. Finally, to measure the α−band instability, we use the total variation (see Eq.20 below).

**Figure 2:**
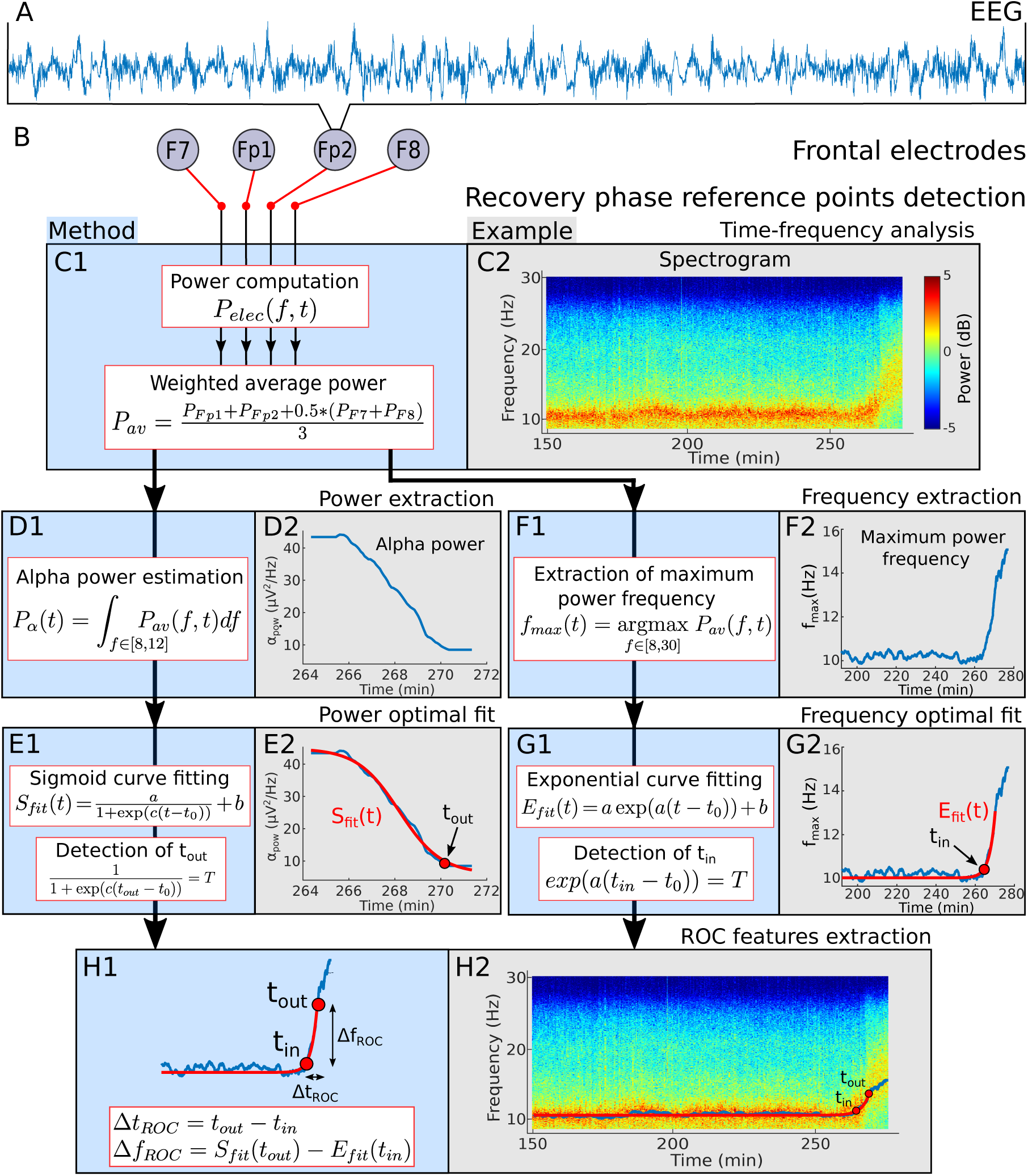
Recovery phase automated detection procedure. **(A)** EEG signal. **(B)** Frontal electrodes. **(C) 1-** Power computation of a weighted average of the four EEG channels. **2-** Spectrogram visualization for a single patient during general anesthesia. **(D) 1-** Estimation of the *α*−band power *P*_*α*_ **2-** Power *P*_*α*_ computed from the spectrogram during [264-272] min corresponding to the end of anesthesia. **(E) 1-** Sigmoid function *S*_*fit*_ used to fit the power *P*_*α*_ during the recovery phase. Definition of the final time *t*_*out*_ for which the fit *S*_*fit*_(*t*_*out*_) = *aT* + *b* reaches for the first time the threshold *aT* + *b*. **2-** Example of a fit and detection of the time *t*_*out*_.**(F) 1-** Estimation of the maximum power frequency *f*_*max*_. **2-** Extracted *f*_*max*_ from the spectrogram. **(G) 1-** Exponential function *E*_*fit*_ to fit *f*_*max*_ and definition of the time *t*_*in*_ where the frequency shift begins to increase. **2-** Extracted *f*_*max*_ and time *t*_*in*_. **(H)1-**Definition of the duration and frequency shifts during recovery **2-** Positioning of the two time points *t*_*in*_ and *t*_*out*_ on the associated spectrogram.

### 3.1 Segmentation of the recovery phase features from the EEG

To quantify the time and spectral changes occurring in the EEG during the recovery phase, we decided to extract the maximum power frequency within the *α*−band by introducing two reference time points *t*_*in*_ and *t*_*out*_. The first time *t*_*in*_ is defined as the first instant where the *α*−band starts to deviate from the steady-state regime which usually characterizes the maintenance phase. The time *t*_*out*_ corresponds to the instant where the power of the α−band has reached a minimum. To estimate these two reference points, we developed a procedure based on EEG signal. It is divided into several steps:

1. Using Fourier transform on a 20 seconds sliding window, we computed the spectrogram of the EEG signal from the four frontal electrodes *P*_*Fp*1_, *P*_*Fp*2_, *P*_*F*7_ and *P*_*F*8_ (Fig.2A-B-C).
2. To account for the predominant role of the frontal cortex generated in the genesis of the α−band during GA [29, 30], we averaged the spectrograms with different weights: the contribution of electrodes F7 and F8 is divided by two (Fig.2C).
3. The signal power within the *α*−band *P*_*α*_(*t*) is equal to the area under the curve *P*_*av*_(*f, t*) for *f* varying in the *α*−band (Fig.2D):

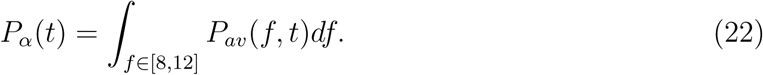 We fitted the power *P*_*α*_(*t*) to a sigmoid curve *S*_*fit*_(*t*) (see Eq.(1)) and defined the time *t*_*out*_ as the first instant for which the curve *S*_*fit*_(*t*) reaches a threshold *T* = 10% (Fig.2E).
4. The frequency curve *f*_*max*_(*t*) with maximal power in the range (8-30) Hz (Fig.2F) is defined by the formula

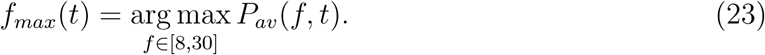 The reference time *t*_*in*_ is then estimated by optimally fitting the function *f*_*max*_(*t*) with an exponential *E*_*fit*_(*t*) = *a* exp(*a*(*t* − *t*_*in*_)) + *b*, where *a >* 0 and *b >* 0 are two parameters to be estimated (see Eq.4) and computing the first time where the function *E*_*fit*_(*t*) deviates from the steady-state *b* by a frequency shift of 0.5 Hz (see Methods and Fig.2G).
5. Once the two reference points *t*_*in*_ and *t*_*out*_ are estimated, we defined the duration and the frequency shifts associated with the *α*−band increase and disappearance (see Fig.2H)

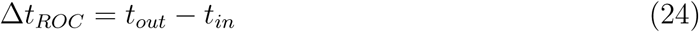

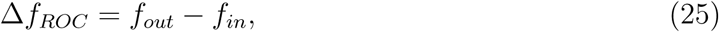

### 3.2 *α*−band and IES collected from induction and maintenance are not correlated with recovery

#### 3.2.1 Statistical correlations for children

To determine whether we could predict the statistical properties of the recovery phase, we first examined possible correlations from the different variables that we extracted from the EEG: we thus examined the parameters associated to IES for children. We study the total time spent in IES during Maintenance and the IES duration induced by a propofol bolus at the end of Induction (Fig.3A-D).

**Figure 3:**
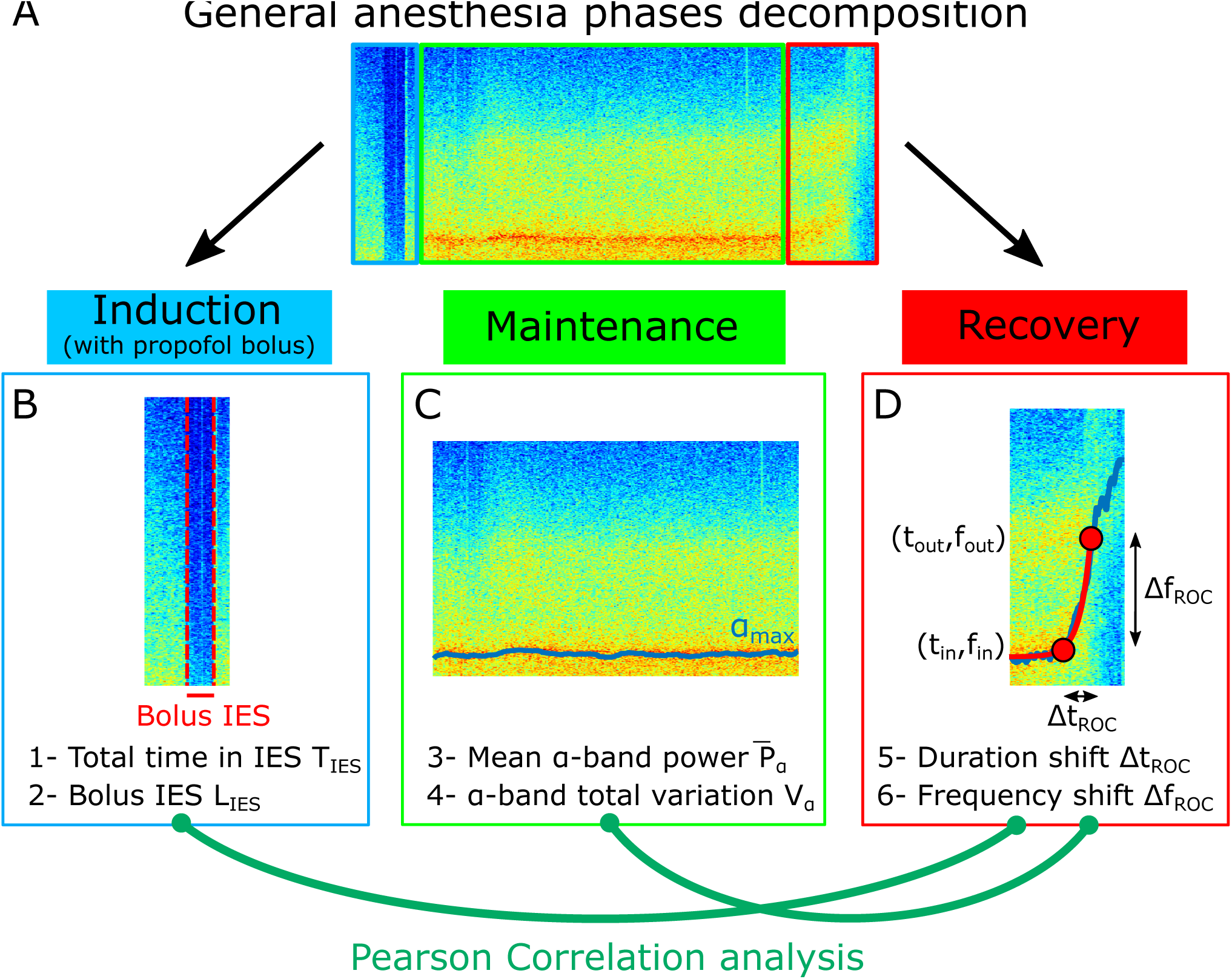
Pearson correlation analysis of the three phases of anesthesia. **(A)** Spectrogram (Time-frequency) representation of the EEG signal during the three phases: induction (blue), maintenance (green), recovery (red). **(B)** Statistical parameters collected during induction: 1- Total time in IES *S*_*IES*_, 2- Longest IES duration *L*_*IES*_. **(C)** Statistical parameters collected during maintenance: 3- Mean *α*−band power 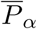, 4- *α*−band total variation *V*_*α*_ **(D)** Statistical parameters collected during recovery: 5- Duration shift Δ*t*_*ROC*_, 6- Frequency shift Δ*f*_*ROC*_).

Using the procedure described in subsection 2.3, we first extracted the duration shift of the recovery phase Δ*t*_*ROC*_ = 7.3 [4.8, 12.7] min (median [IQR]), and the frequency shift Δ*f*_*ROC*_ = 3.5 [2.3, 5.4] Hz, for children. We also extracted (see subsection 2.5) the total time spent in IES *S*_*IES*_ = 1.3 [0.5, 2.1] min and the longest time spent in IES (induced by a propofol bolus) *L*_*IES*_ = 0.3 [0.1, 0.9] min. Secondly, we computed the Pearson correlation coefficients and found that the total duration of the IES (resp. longest IES) is not correlated either to the duration of recovery Δ*t*_*ROC*_ (Pearson, *r* = 0.20; p-value, *p* = 0.15 (resp. *r* = 0.08; *p* = 0.58)) or to the shift frequency Δ*f*_*ROC*_ (*r* = 0.03; *p* = 0.86 (resp. *r* = −0.21; *p* = 0.14)), as shown in Fig.5B1-4. At this stage, we conclude that there are no significant correlation revealed by the transient parameters associated with the absence of the EEG signal.

To further study the possible correlations between phases, we extracted other statistics by computing the mean power 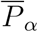 of the *α*−band during the maintenance and the total variation *V*_*α*_ that measures the deviation of the maximum frequency *f*_*max*_ of the α−band from a flat curve (Fig.3C-D). We report the mean power 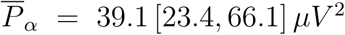 and *V*_*α*_ = 0.7 [0.4, 1.3] *Hz/min*. We estimated the correlations between 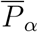 (resp. *V*_*α*_) and either Δ*t*_*ROC*_ (*r* = 0.11; *p* = 0.46 (resp. *r* = 0.07; *p* = 0.58)) or Δ*f*_*ROC*_ (*r* = −0.01; *p* = 0.95 (resp. *r* = 0.18; *p* = 0.21)) for the children group. (Fig.5B5-8) and found no significant correlations. To conclude the statistics associated with the presence or absence of the α−band do not show any significant correlation with the recovery dynamics.

#### 3.2.2 Statistical correlations for adults

We applied the same procedure for adults, sedated with propofol injected with a target controlled infusion (TCI) protocol. We found that the duration shift of the recovery phase was Δ*t*_*ROC*_ = 10.4 [4.8, 12.3] min, and the frequency shift Δ*f*_*ROC*_ = 2.1 [1.5, 3.3] Hz, *S*_*IES*_ = 0.1 [0, 1.6] min and *L*_*IES*_ = 0.06 [0, 0.1] min. We found that the total time in IES (resp. longest IES) was not correlated to either the duration shift Δ*t*_*ROC*_ (*r* = 0.13; *p* = 0.46 (resp. *r* = −0.05; *p* = 0.78)) or the frequency shift Δ*f*_*ROC*_ (*r* = 0.03; *p* = 0.87 (resp. *r* = −0.02; *p* = 0.91)) (Fig.4B1-4). We find that the mean power 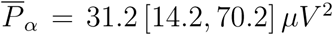 and *V*_*α*_ = 1.0 [0.8, 1.6] Hz. Finally, we found no significant correlations between the power 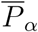 (resp. *V*_*α*_) and either Δ*t*_*ROC*_ (*r* = 0.13; *p* = 0.45 (resp. *r* = 0.05; *p* = 0.76)) or Δ*f*_*ROC*_ (*r* = 0.01; *p* = 0.95 (resp. *r* = −0.09; *p* = 0.62)), as shown in Fig.4B5-8.

**Figure 4:**
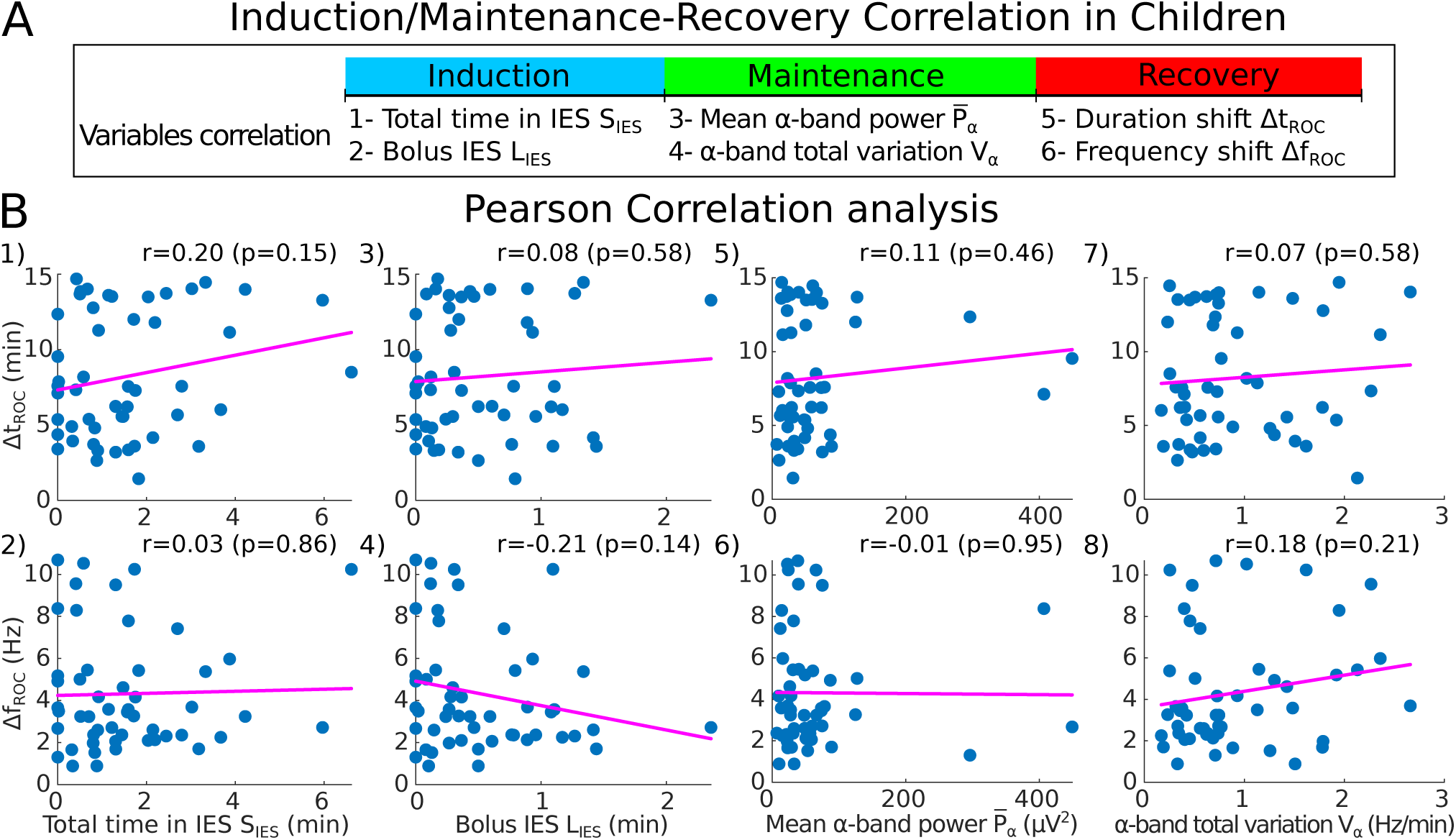
Pearson correlation analysis between maintenance vs recovery and induction vs recovery for 51 children. **(A)** Summary of the statistical parameters collected from the induction and maintenance phases (total time in IES *S*_*IES*_, bolus IES duration *L*_*IES*_, mean *α*−band power 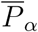 and *α*−band total variation *V*_*α*_) and the recovery phase markers (duration shift Δ*t*_*ROC*_ and frequency shift Δ*f*_*ROC*_). **(B)** Scatter plots and Pearson correlation (coefficient *r*, p-value *p*) between Δ*t*_*ROC*_ (Δ*f*_*ROC*_) and 1-2) *S*_*IES*_, 3-4) *L*_*IES*_, 5-6) 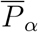 and 7-8) *V*_*α*_.

**Figure 5:**
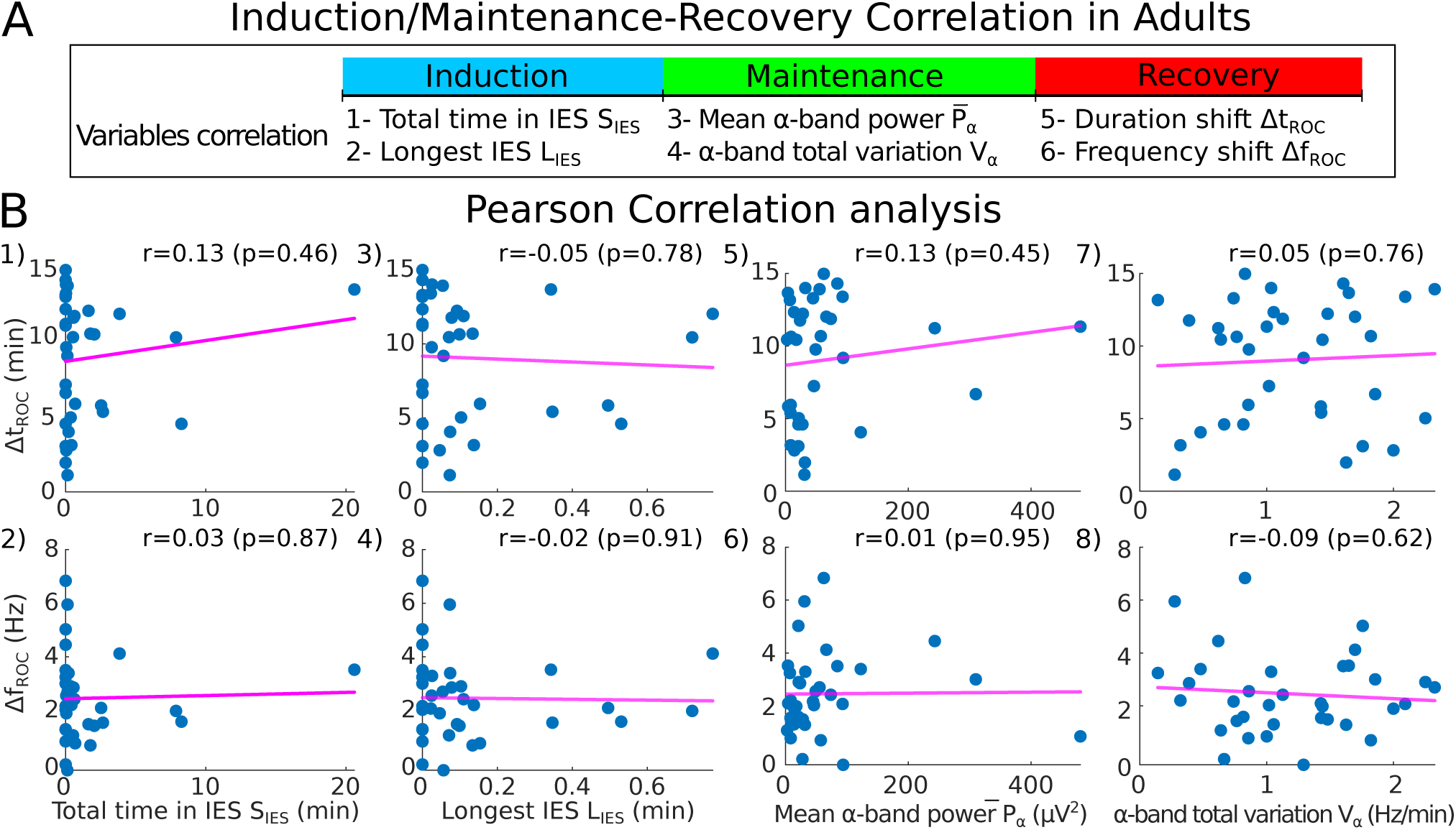
Pearson correlation analysis between maintenance vs recovery and induction vs recovery for 35 adults. **(A)** Summary of the statistical parameters collected during induction and maintenance phases (*S*_*IES*_, *L*_*IES*_, 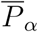 and *V*_*α*_) and the recovery phase(Δ*t*_*ROC*_ and Δ*f*_*ROC*_). **(B)** Scatter plots and Pearson correlation (coefficient *r*, p-value *p*) between Δ*t*_*ROC*_ (Δ*f*_*ROC*_) and 1-2) *S*_*IES*_, 3-4) *L*_*IES*_, 5-6) 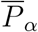 and 7-8) *V*_*α*_.

We conclude using a statistical approach estimated over two cohorts of adult and children that the duration of the recovery phase, characterized by a systematic shift of the maximum *α*−band is not correlated neither with the fraction of iso-electric suppression nor the *α* band dynamics measured by its power and variation. Thus, these EEG parameters obtained during induction and maintenance of GA cannot be used to predict the duration of emergence from the *α*−band to the *β* rhythms.

## 4 Discussion

In the present study, we aimed at searching for possible correlations between the EEG features of different phases of general anesthesia. For that goal, we used several mathematical indicators computed in each phase. During induction, we considered the longest IES (induced by a bolus of propofol in children). While adults underwent intravenous hypnotic administration, children underwent GA induction through inhalation of sevoflurane followed by a bolus of propofol to avoid oro-tracheal intubation complications [31]. During maintenance, we computed the mean power 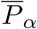 of the *α*−band and the total variation *V*_*α*_ (Eq.(21)), a measure of the *α*−oscillations maximum amplitude instability. A small value for *V*_*α*_ represents low fluctuations around a flat line. Consequently, the higher is *V*_*α*_, the larger is the deviation. During both the induction and maintenance phases, we estimated the total time spent in IES. Finally, during the recovery phase, we estimated the frequency and the duration shifts associated to the α−band disappearance (Fig.2). The present analysis revealed no correlations between these parameters (Fig.6). These results invalidate our hypothesis concerning the possible causality between longest IES or large α−band dynamics which could have been correlated with a long duration to recover consciousness from a dominant α−band. These results suggest that IES durations and α−band dynamics cannot be used as predictive indicators of recovery from anesthesia.

**Figure 6:**
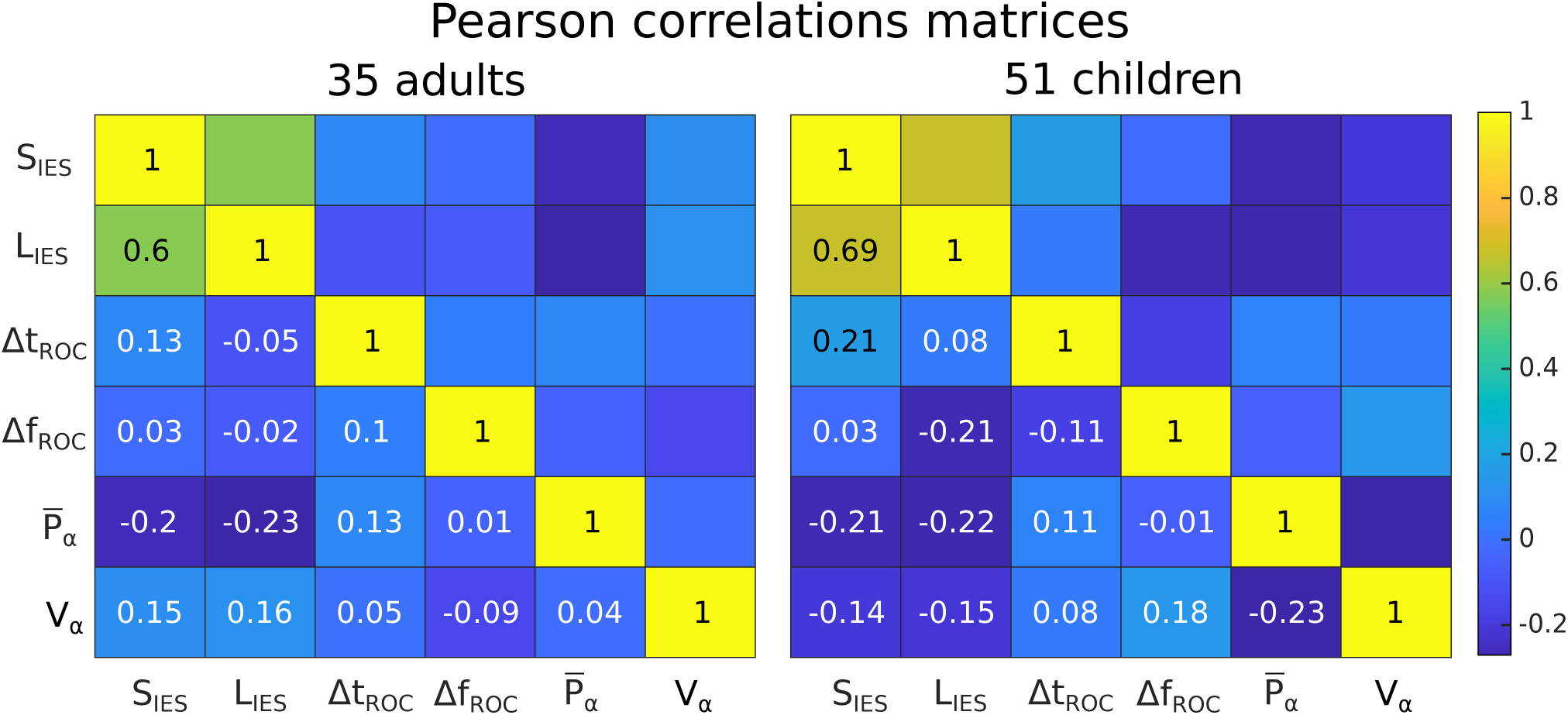
Pearson correlation heatmaps obtained separately for adults and children.

IES duration are not only uncorrelated to the recovery duration, defined as the disappearance of the dominant *α*−band in the EEG time-frequency representation [29] (Fig.3), but also to the degree of cognitive impairment after anesthesia [24], where the recovery was computed differently by fitting the decay of expired isoflurane to a single exponential. By defining the clinical recovery duration from the instant where the hypnotic injection/inhalation is shut down to the extubation time, where the patient showed first signs of awareness or recovery of effective spontaneous ventilation. We confirmed here the hypothesis that there are no correlation between IES and the recovery durations as shown in Fig.7.

**Figure 7:**
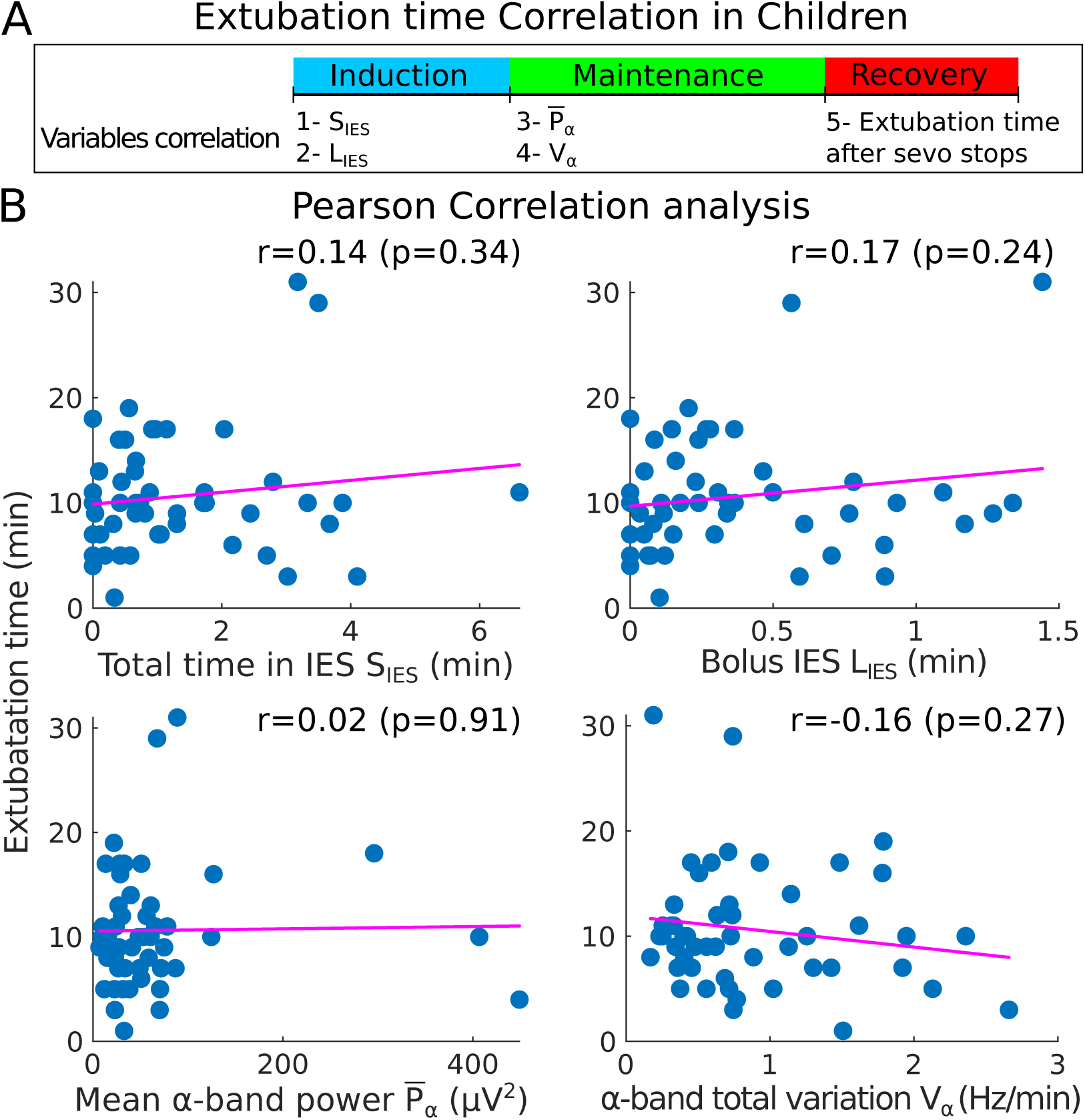
Pearson correlation between the induction/maintenance phases and the extubation time for the children group. **(A)** Statistical parameters collected during induction and maintenance (*S*_*IES*_, *L*_*IES*_, 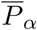 and *V*_*α*_) and the extubation time after the administration of sevoflurane is stopped. **(B)** Scatter plots and Pearson correlation (coefficient *r*, p-value *p*) between the extubation time and 1- *S*_*IES*_ (top left), 2- 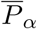 (bottom left), 3- *L*_*IES*_ (top right) and 4- *V*_*α*_ (bottom right).

In addition, the *α*−band dynamics did not show further correlations as revealed by the total variation (eq. 21) of the α−band maximum frequency *α*_*max*_(*t*). A high total variation could reflect a constant re-adjustment that could be induced by a real time hypnotic concentration variation [32, 33]. Interestingly, we reported a significant correlation between the time spent in α−suppressions, defined as a transient frequency loss of the α−rhythms in the EEG signal, and the total variation (Pearson *r* = 0.38, p-value *p* = 0.02) (Fig.8). This result suggests that high oscillations of the maximum amplitude of the α−band are associated with a loss of the band and thus reveal a possible instability of the brain during general anesthesia. Possibly, the total variation *V*_*α*_ could anticipate the appearance of the α−suppression that precedes the occurrence of IES. The total variation *V*_*α*_ could either increase due to a change in the hypnotic concentration or due to the intrinsic response of the brain to a fixed concentration. In the latter case, these changes could represent an instability of thalamo-cortical neuronal networks to converge to a stable constant α−oscillation, which is a marker of brain synchronization [8]. Moreover, the mean *α*−band power 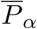 has been used to monitor the depth of anesthesia and as a possible predictor for the arrival of IES [14, 34]. Lower values of the power 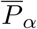 reflects a vulnerable brain and could have been associated to a longer time of emergence. However, this hypothesis is not confirmed in our analysis (Fig.7 bottom), where there are no significant correlations between the 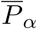 and the ROC frequency and time shifts. Interestingly, we found that the distribution of Δ*t*_*ROC*_ is similar for both adults and children groups (Table 1). Finally, the ROC reflects the transient recovery of the brain from a constant injection of anesthetic. In the case of GABAergic product [35], that increases the strength of the inhibitory neurons, the dominant neuronal activity is driven by the speed at which the product is eliminated by the body. This mechanism is probably completely different from the anesthesia induced steady-state. Indeed, thalamic neurons are intrinsically changing states under general anesthesia [36], switching from spiking to bursting during loss of consciousness, ROC should be associated to the reverse switching (bursting to fast spiking). This switching is associated with the loss in the delta (0.1-4 Hz) power and the increase of beta (12-30 Hz) power as the neuronal activity recovers to its normal state [29].

**Figure 8:**
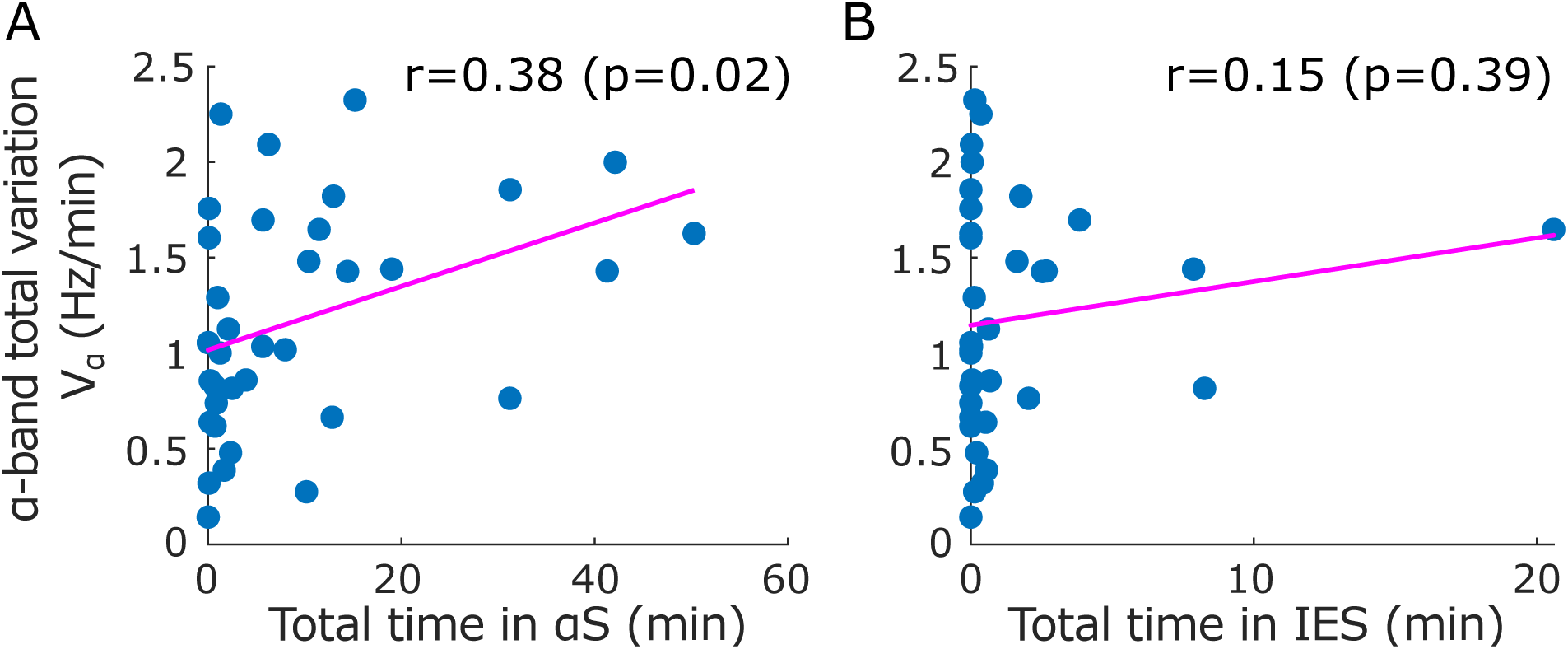
Correlation between the time spent in suppressions and the total variation of the *α*−band computed for the adults population. Scatter plots and Pearson correlation (coefficient *r*, p-value *p*) between the *α*−band total variation *V*_*α*_ and **(A)** the total time spent in *α*−suppressions (frequency loss in the *α*−band), **(B)** the total time spent in IES *S*_*IES*_.

**Table 1:** Medians, inter-quartile ranges and Pearson’s correlations of EEG statistical features

| Measure | Median | Q1 | Q3 | Pearson's correlation |  |
| --- | --- | --- | --- | --- | --- |
|  |  |  |  | 1. | 2. |
| **Adults** |  |  |  |  |  |
| 1. $\Delta t_{ROC}$ (min) | 10.4 | 4.8 | 12.3 | | |
| 2. $\Delta f_{ROC}$ (Hz) | 2.1 | 1.5 | 3.3 | | |
| 3. $S_{IES}$ (min) | 0.1 | 0 | 1.6 | 0.13 | 0.03 |
| 4. $L_{IES}$ (min) | 0.06 | 0 | 0.1 | −0.05 | −0.02 |
| 5. $\overline{P}_\alpha$ ($\mu V^2$) | 31.2 | 14.2 | 70.2 | 0.13 | 0.01 |
| 6. $V_\alpha$ (Hz/min) | 1.0 | 0.8 | 1.6 | 0.05 | −0.09 |
| **Children** |  |  |  |  |  |
| 1. $\Delta t_{ROC}$ | 7.3 | 4.8 | 12.7 | | |
| 2. $\Delta f_{ROC}$ | 3.5 | 2.3 | 5.4 | | |
| 3. $S_{IES}$ | 1.3 | 0.5 | 2.1 | 0.20 | 0.03 |
| 4. $L_{IES}$ | 0.3 | 0.1 | 0.9 | 0.08 | −0.21 |
| 5. $\overline{P}_\alpha$ | 39.1 | 23.4 | 66.1 | 0.11 | −0.01 |
| 6. $V_\alpha$ | 0.7 | 0.4 | 1.3 | 0.07 | 0.18 |

Notes: $*p < .05$

### General statement

This prospective observational study included patients receiving a scheduled elective procedure requiring general anesthesia. The EEG data used in the study were recorded from Louis-Mourier Hospital (Colombes, France), collected between October 2019 and September 2021 and included 51 children and 35 adults. Age group and clinical information were also provided. For adult patients, general anesthesia was induced by intravenous administration of a opioid agent (remifentanil or sufentanil) and a hypnotic agent (propofol), the doses of which were defined according to the Minto and Schnider models [25].

This study is followed by the Institutional Review Board of La Société de Réanimation. In agreement with the ethics committee for this non-interventional study an information letter was given and an oral agreement was obtained from each patient before anesthesia. The protocol was approved by Louis-Mourier Hospital (Colombes, France) following the guideline of the ethics committee, and therefore, has been performed in accordance with the ethical standards laid down in 1964 declaration of Helsinki and its later amendments.

For children, the hypnotic agent (sevoflurane) was administrated by inhalation and followed by one or more bolus of propofol until the appearance of iso-electric suppressions for oro-tracheal intubation. The initial quantity of propofol was calibrated according to the children weight. Depending on the patient reaction and the anesthesia phase, the anesthesiologist can adjust the concentrations as long as standard of care is respected.

## Data Availability

All data produced in the present study are available upon reasonable request to the authors

## Data availability

The datasets generated and analysed in the current study are available from the corresponding authors upon reasonable request.

## Acknowledgment

D.H. research is supported by a grant ANR NEUC-0001, PSL and CNRS pre-maturation and the European Research Council (ERC) under the European Unions Horizon 2020 research and innovation programme (grant agreement No 882673).

## Abbreviations

EEG: ElectroEncephaloGram
GA: General Anesthesia
IES: Iso-Electric Suppression
ROC: Recovery Of Consciousness

